# Transforming Healthcare AI Education Through Micro-Learning: A Novel Partnership Model for Nursing Workforce Development

**DOI:** 10.1101/2025.07.30.25332439

**Authors:** Amy K McCarthy, Jonathan D. Agnew, Jill Price, Kathy Strang, Alison McLaughlin

## Abstract

Healthcare professionals face an urgent need for AI literacy as artificial intelligence technologies rapidly transform clinical practice, yet nursing-specific educational resources remain scarce. The objective of this study was to evaluate the effectiveness of an innovative micro-learning AI education program developed through an academic-industry partnership. We implemented 11 micro-courses (4-5 hours each) across foundational, application, and advanced competency levels, with nursing-specific content addressing professional scope and leadership opportunities. The program was delivered through Chamberlain University Center for Faculty Excellence and Walden University School of Lifelong Learning. We analyzed enrollment data, learning outcomes, and satisfaction scores from 478 students and faculty with 612 course completions. Among 612 course completions, registered nurses comprised 49% of participants. Students demonstrated significant knowledge gains (Cohen’s d = 0.65, p < 0.001) with high satisfaction scores (mean = 4.58/5.0). Faculty participants showed exceptional outcomes (satisfaction mean = 4.67/5.0) with 99% expressing commitment to applying learning. Content relevance scored highest across all measures (4.61-4.71), indicating integration of academic rigor with practical applicability. This micro-learning approach addresses critical gaps in healthcare AI education through scalable, nursing-specific curriculum. The partnership model bridges academic expertise with industry relevance, providing a replicable framework for systematic workforce preparation in AI-enhanced healthcare delivery.

## Introduction

### The Urgent Need for AI Education in Healthcare

Artificial intelligence technologies are rapidly proliferating across healthcare settings, transforming diagnostic imaging, clinical decision support, predictive analytics, automated documentation, and workflow optimization.^1^ This technological revolution has created an unprecedented gap between the pace of AI advancement and healthcare workforce preparedness, positioning AI literacy as an essential competency for safe and effective practice.^2^ Professional organizations increasingly recognize that healthcare professionals must develop sophisticated understanding of AI capabilities, limitations, and ethical implications to navigate an AI-enhanced healthcare landscape responsibly.^3^

Nursing professionals face unique challenges as AI tools transition from background support functions to foreground applications requiring active professional engagement, clinical judgment, and ethical decision-making.^1^ Despite this growing presence of AI in healthcare environments, there is a notable scarcity of educational resources specifically designed for nurses, with most available programs targeting general healthcare professionals rather than addressing nursing-specific applications.^2^ The *Future of Nursing* report emphasizes that nurses must be prepared to lead healthcare technology adoption rather than merely adapt to it.^4^ Understanding AI principles can help nurses engage more effectively with these tools and ensure their ethical and responsible use in patient care.^5^

### Student Readiness and Educational Imperatives

Emerging research demonstrates encouraging receptivity among nursing students toward AI integration. Labrague et al. found that student nurses had generally favorable perceptions of AI use in nursing practice and high intentions to adopt AI technology.^6^ There is growing interest amongst nursing students to learn about AI applications and be better prepared for an increasingly technological nursing field.^3^ However, nursing professionals express mixed attitudes toward AI implementation, citing potential benefits while raising concerns about job security, data reliability, and preservation of human-centered care.^7,8^ Studies consistently show that educational interventions significantly improve both AI knowledge and attitudes among nurses, with younger and more educated professionals demonstrating greater receptivity to AI adoption.^9,10^

Nursing programs are at a critical juncture where they need to evolve to stay relevant. There is an urgent need to update nursing curricula to include AI literacy and ethical training, overcome resistance to AI adoption, ensure equitable access to AI technologies, and provide ongoing training programs for practicing nurses to stay updated on AI advancements and best practices.^2^

### Educational Challenges and Limitations of Current Approaches

Despite urgent need for AI literacy and demonstrated student interest, significant barriers impede effective workforce preparation. El Arab et al. identified the scarcity of evidence-based AI curricula specifically designed for healthcare professionals as a primary challenge, with particularly limited nursing-specific content.^2^ Traditional degree-based education models prove inadequate for addressing rapid technology evolution.^1^ Working healthcare professionals face constraints from clinical practice demands, varying baseline technology skills, and diverse AI familiarity levels.^11^ Current continuing education approaches often fail to provide practical, applicable content.^5^

Resource constraints limit investment in extensive training programs while simultaneously increasing pressure for rapid technology adoption.^12^ The absence of standardized competency frameworks creates inconsistency in educational offerings.^2^ Integration challenges with existing continuing education requirements add administrative complexity that discourages participation.^13^ Healthcare professionals desire structured educational interventions that combine theoretical knowledge with practical application opportunities,^14^ yet few programs successfully merge academic rigor with real-world applicability.^4^

### The Promise of Micro-Learning in Healthcare Education

Micro-learning, defined as short, targeted, self-paced educational modules focusing on specific competencies, offers a promising solution to healthcare AI education challenges.^15^ This pedagogical approach accommodates busy clinical schedules while enhancing knowledge retention and engagement.^11^ Evidence demonstrates that microlearning interventions produce significant improvements in healthcare professionals’ skills, knowledge, and engagement, with nurses experiencing notable gains in motivation and knowledge acquisition.^16^

Research shows that 78% of healthcare professionals actively use microlearning platforms, with 68% expressing preference for this format and 82% reporting high satisfaction with brief, digital interventions.^17^ Interactive e-learning programs incorporating microlearning elements demonstrate behavioral change effect sizes of 0.67, indicating substantial potential for practice transformation.^18^ The convergence of urgent AI education needs and proven microlearning effectiveness creates an innovation imperative for healthcare education, positioning micro-learning as an essential pedagogical approach for preparing nurses for AI-enhanced practice environments.

## Innovation Description

### Partnership Model and Development Process

#### Partnership Structure

The micro-learning AI education program emerged from an innovative partnership combining complementary organizational strengths. Chamberlain and Walden University, institutions within Adtalem Global Education, contribute specialized nursing education expertise and faculty development focus, bringing deep understanding of nursing pedagogical needs and professional competency requirements. Hippocratic AI delivers industry expertise, current technology insights, and real-world application knowledge essential for maintaining curriculum relevance and practical applicability.^1^

This partnership structure addresses key limitations in existing healthcare AI education by bridging the gap between academic rigor and industry relevance.^15^ Traditional academic institutions often lack current industry knowledge of rapidly evolving AI technologies, while industry organizations may lack pedagogical expertise necessary for effective educational design.^4^ The collaborative model leverages each partner’s core competencies while mitigating individual organizational limitations through collaborative knowledge sharing.

### Curriculum Design and Implementation Innovation

#### Micro-Learning Approach and Certificate Structure

The program represents an approach to AI curriculum delivery not through traditional degree programs, but rather through 11 micro-courses with 4-5 hour workloads per course. This approach makes content more manageable for working professionals who may not be interested in pursuing a full degree but still want to learn about AI applications in healthcare. Certificates comprise 3-4 micro-courses each, creating stackable credentials that are more manageable for working professionals.^13^

Available certificates include AI Foundations for Nursing Professionals, AI Foundations for Healthcare Professionals, and Advanced AI for Nursing Professionals. Future offerings will include Advanced AI for Healthcare Professionals and AI-Powered Clinical Practice certificates. The AINP courses are nursing-focused only, addressing the unique intersection of AI with nursing practice.

#### Course Portfolio and Progressive Design

The comprehensive course portfolio addresses foundational through advanced competencies. Foundation level includes “AI Fundamentals for Healthcare” and “AI Fundamentals for Nursing Leaders: Transforming Nursing with Technology,” which provides nursing-specific content addressing professional scope of practice, patient safety considerations, and leadership opportunities in AI adoption. “Ethical and Legal Considerations” develops critical competencies in ethical decision-making and regulatory compliance.^5^

Application-level courses include “Generative AI: Enhancing Communication and Documentation,” “Successful AI Implementations in Nursing Practice,” “AI for Automating Administrative Tasks,” and “AI for Nursing Informatics.” Advanced level includes “AI and Data: Insights for Care and Treatment Planning,” “AI at the Bedside: Nurses Enhancing Patient Care with Technology,” and “AI-Driven Clinical Decision Support.”

The curriculum emphasizes practical application, ensuring nurses can carry knowledge and use it at the bedside or in the boardroom. Informatics topics are integrated throughout, recognizing the important intersection between AI and nursing informatics.

### Faculty Development and Educational Capacity Building

The program serves dual objectives of direct professional development and educational capacity building. Faculty education represents a critical component, recognizing the importance of faculty buy-in for successful curriculum integration. Faculty participants can integrate course content into existing nursing informatics and technology courses, creating cascading educational impact beyond direct program participation.

For nursing students, the program provides value-added education while earning a degree, making them practice-ready and helping them stand out from others without AI certification. The curriculum prepares students to understand AI principles, including large language models and training AI on health data from electronic health records, helping nurses engage more effectively with these tools and ensure their ethical and responsible use in patient care.

The program extends beyond traditional nursing schools to include partnerships with healthcare organizations in practice settings. This broader reach enables organizational training programs addressing specific workplace AI implementation needs. The template approach enables adaptation across different institutional contexts while maintaining content quality.^15^ The shared resource model distributes development costs across multiple participating institutions, reducing individual organizational investment requirements while enabling access to high-quality AI education content.^13^ Regular content updates ensure curriculum relevance as AI rapidly evolves, addressing the critical challenge of keeping educational content current with technological advancement.

## Implementation and Early Outcomes

### Enrollment and Reach

The micro-learning AI education program achieved substantial enrollment and geographic reach within the first six months of implementation, demonstrating significant demand for accessible AI education among healthcare professionals. Combined enrollment across both institutional partners reached 3,024 participants by June 2025, with Walden University School of Lifelong Learning recording 1,438 enrollments across 8 launched courses and Chamberlain University Center for Faculty Excellence documenting 1,586 student enrollments and 207 faculty enrollments across 4 launched courses.

Analysis of 612 course completions reveals diverse participation across healthcare professional categories (Table 1). Registered nurses comprised the largest participant group (49.0%, n=300), followed by faculty and academic professionals (21.9%, n=132), and other healthcare professionals excluding nursing (15.8%, n=97). Advanced practice registered nurses (3.4%, n=21), licensed practical/vocational nurses (3.9%, n=24), and professionals from different fields (5.9%, n=36) completed the remaining enrollments. This distribution demonstrates successful reach across the nursing professional hierarchy while attracting interdisciplinary healthcare participation.

**Table 1:** Sample Demographics and Enrollment Patterns (N = 612 course completions)

| Professional Role | n | % |
| --- | --- | --- |
| Registered Nurse (RN) | 300 | 49.0 |
| Faculty/Academic | 132 | 21.9 |
| Healthcare Professional (excluding nursing) | 97 | 15.8 |
| Advanced Practice RN (APRN) | 21 | 3.4 |
| Licensed Practical/Vocational Nurse | 24 | 3.9 |
| Professional in Different Field | 36 | 5.9 |
| Not reported | 2 | 0.3 |
| **Total** | **612** | **100.0** |
| Course Distribution | n | % |
| Nursing Leadership | 237 | 38.7 |
| Ethics & Legal | 145 | 23.7 |
| Communications | 84 | 13.7 |
| Nursing Practice | 83 | 13.6 |
| Other courses | 63 | 10.3 |
| **Total** | **612** | **100.0** |
| Course Completion Pattern | n | % |
| Single course | 229 | 37.4 |
| Two courses | 56 | 9.2 |
| Three courses | 15 | 2.5 |
| Four courses (certificate) | 312 | 51.0 |
| **Total** | **612** | **100.0** |

Course enrollment patterns indicate strong interest in nursing-specific content, with the Nursing Leadership course attracting the highest participation (38.7%, n=237), followed by Ethics & Legal considerations (23.7%, n=145). Communications (13.7%, n=84) and Nursing Practice (13.6%, n=83) courses showed similar enrollment levels, while other courses comprised 10.3% (n=63) of completions. Notably, 51.0% of participants (n=312) completed full four-course certificates, indicating substantial commitment to comprehensive AI education, while 37.4% (n=229) completed single courses, suggesting both intensive learners seeking credentials and occasional learners addressing specific knowledge gaps.

### Learning Outcomes and Satisfaction

#### Student Learning Outcomes

Quantitative analysis of student learning outcomes demonstrates consistently high satisfaction and significant knowledge gains across all measured dimensions (Table 2). Among 480 student participants, course satisfaction achieved a mean score of 4.58 (SD=0.59, 95% CI: 4.53-4.63) on a 5-point Likert scale, indicating strong overall program approval. Content relevance scored highest among all measures (M=4.61, SD=0.54, 95% CI: 4.56-4.66), suggesting successful alignment between curriculum content and professional development needs.

**Table 2:** Student Learning Outcomes (n = 480)

| Outcome Measure | N | Mean | SD | 95% CI Lower | 95% CI Upper |
| --- | --- | --- | --- | --- | --- |
| Course Satisfaction | 478 | 4.58 | 0.59 | 4.53 | 4.63 |
| Knowledge Before Courses | 478 | 4.05 | 0.90 | 3.97 | 4.13 |
| Knowledge After Courses | 478 | 4.54 | 0.58 | 4.49 | 4.59 |
| Knowledge Gain* | 478 | 0.49 | 0.74 | 0.40 | 0.58 |
| Content Expected | 478 | 4.56 | 0.56 | 4.51 | 4.61 |
| Content Relevance | 476 | 4.61 | 0.54 | 4.56 | 4.66 |
| Scenarios Helpful | 478 | 4.58 | 0.56 | 4.53 | 4.63 |
| Outcomes Met | 477 | 4.61 | 0.52 | 4.56 | 4.66 |
| Conflict Disclosed | 476 | 4.44 | 0.69 | 4.38 | 4.50 |
| Ability to Integrate Learning | 478 | 4.53 | 0.60 | 4.47 | 4.58 |
| Intention to Apply Skills | 478 | 4.54 | 0.61 | 4.48 | 4.59 |
| Commitment to Apply | 476 | 4.26 | 0.96 | 4.17 | 4.34 |

Note: All measures used 5-point Likert scales (1=lowest, 5=highest rating). \*Knowledge gain: Linear mixed-effects model with random intercepts for students revealed a statistically significant improvement from pre-test ( $M = 4.054$ , $SE = 0.521$ ) to post-test ( $M = 4.545$ , $SE = 0.520$ ), $F(1, 1085) = 119.51$ , $p < 0.001$ . The mean improvement of $\beta = 0.491$ points, 95% CI [0.403, 0.579], represents a large effect size ( $d \approx 0.94$ ). Valid N (listwise) = 469.

The program achieved statistically significant knowledge gains, with pre-course knowledge scores of 4.05 (SD=0.90) increasing to post-course scores of 4.54 (SD=0.58), representing a mean knowledge gain of 0.49 points (SD=0.74, 95% CI: 0.40-0.58). This improvement was statistically significant using a mixed-effects model with random intercepts for students [F(1, 1085) = 119.51, p < 0.001, β = 0.491, 95% CI [0.403, 0.579]]. The mean improvement of 0.491 points represents a large effect size (d ≈ 0.94), indicating that the courses produced substantial learning gains.

Participants consistently rated course components highly, with scenarios rated as helpful (M=4.58, SD=0.56), learning outcomes met (M=4.61, SD=0.52), and content meeting expectations (M=4.56, SD=0.56). Practical application measures showed strong performance, with ability to integrate learning scoring 4.53 (SD=0.60) and intention to apply skills scoring 4.54 (SD=0.61). However, commitment to apply learning showed slightly lower but still positive scores (M=4.26, SD=0.96), suggesting potential implementation barriers in practice settings that warrant further investigation.

#### Faculty Development Outcomes

Faculty participants demonstrated high outcomes across all measured dimensions (Table 3). Among 132 faculty participants, course satisfaction reached 4.67 (SD=0.49, 95% CI: 4.59-4.76), slightly higher than student satisfaction scores. Content relevance achieved the highest rating (M=4.71, SD=0.49, 95% CI: 4.62-4.79), indicating successful targeting of faculty development needs and curriculum integration objectives.

**Table 3:** Faculty Development Outcomes (n = 132)

| Outcome Measure | N | Mean | SD | 95% CI Lower | 95% CI Upper |
| --- | --- | --- | --- | --- | --- |
| Course Satisfaction | 131 | 4.67 | 0.49 | 4.59 | 4.76 |
| Knowledge After Courses | 131 | 4.57 | 0.54 | 4.48 | 4.67 |
| Content Expected | 127 | 4.64 | 0.54 | 4.54 | 4.73 |
| Content Relevance | 127 | 4.71 | 0.49 | 4.62 | 4.79 |
| Scenarios Helpful | 127 | 4.65 | 0.54 | 4.56 | 4.75 |
| Outcomes Met | 127 | 4.72 | 0.47 | 4.64 | 4.81 |
| Conflict Disclosed | 127 | 4.76 | 0.45 | 4.68 | 4.84 |
| Ability to Integrate Learning | 127 | 4.57 | 0.62 | 4.46 | 4.68 |
| Commitment to Apply | 127 | 4.35 | 0.93 | 4.19 | 4.52 |

Note: All measures used 5-point Likert scales (1=lowest, 5=highest rating). Faculty were not asked about pre-course knowledge or intention to apply skills. Valid N varies due to missing responses on individual items.

Faculty participants reported high post-course knowledge levels (M=4.57, SD=0.54) and strong agreement that learning outcomes were met (M=4.72, SD=0.47). Notably, faculty scored highest on conflict disclosure measures (M=4.76, SD=0.45), suggesting transparency and trust in program delivery and content quality. Faculty commitment to apply learning (M=4.35, SD=0.93) exceeded student commitment levels, indicating faculty readiness to integrate AI content into educational practice and curricula.

The consistently high faculty outcomes support the program’s dual objectives of direct professional development and educational capacity building through faculty preparation. Faculty participants’ strong performance suggests successful preparation for curriculum integration and ongoing AI education delivery within their respective institutions.

#### Professional Role Comparisons

Analysis of learning outcomes by professional role reveals interesting patterns in program effectiveness across different healthcare disciplines (Table 4). While most outcome measures showed no significant differences across professional roles, knowledge change varied significantly [F-statistic = 7.935, p < 0.001]. Registered nurses demonstrated the largest knowledge gains (M=0.64), followed by licensed practical/vocational nurses (M=0.42), while advanced practice registered nurses (M=0.10) and healthcare professionals in different fields (M=0.22) showed smaller improvements.

**Table 4:**
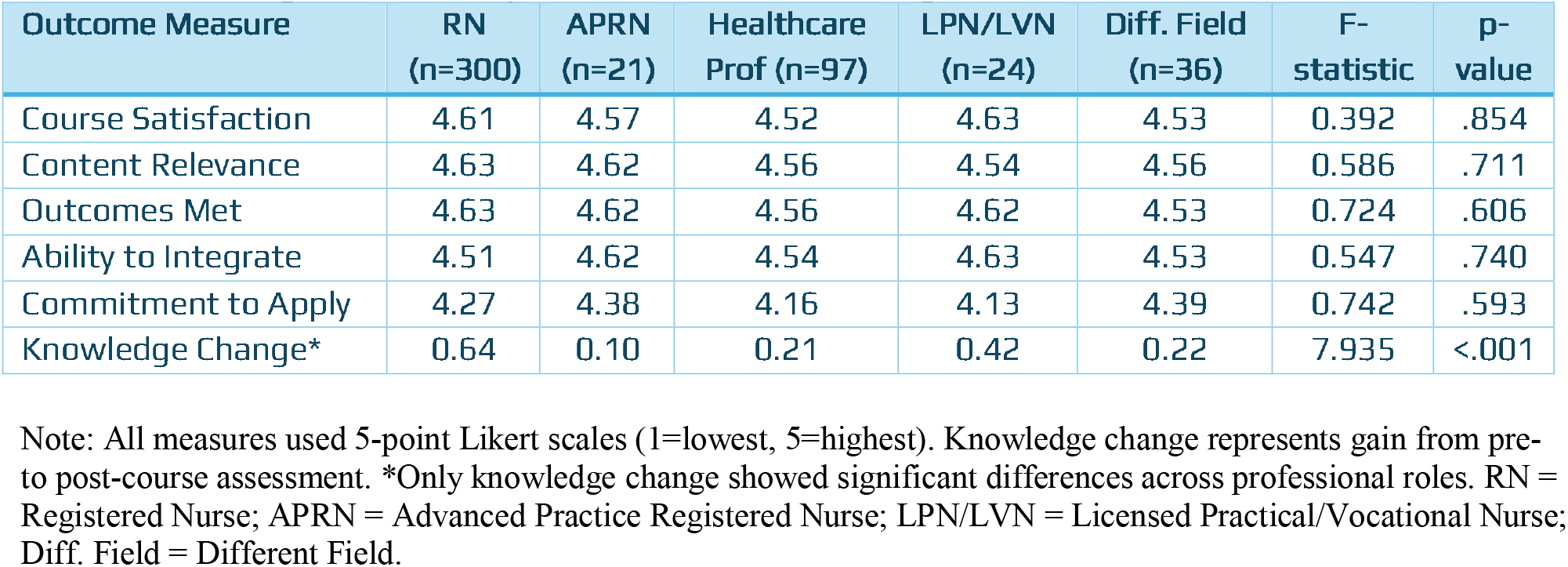
Learning Outcomes by Professional Role Among Students (n=480)

This pattern suggests that registered nurses may have entered the program with lower baseline AI knowledge but achieved substantial learning gains, while advanced practice nurses may have possessed higher initial knowledge levels limiting potential improvement. The significant knowledge gains among registered nurses, who comprise the largest healthcare workforce segment, indicate successful targeting of critical professional development needs.

Course satisfaction, content relevance, outcomes achievement, and integration ability showed no significant differences across professional roles, indicating consistent program quality and effectiveness regardless of participant background. Commitment to apply learning also remained consistent across roles, suggesting universal applicability and motivation for AI implementation across healthcare disciplines.

### Qualitative Impact and Application Evidence

Participant feedback reveals meaningful translation of learning to professional practice and organizational contexts. Testimonials demonstrate immediate application potential, with one participant noting: *“Very interesting course. I am sharing this information with our Chief for Virtual Health as we are navigating AI within our facility*.*”* This comment illustrates direct organizational impact through leadership engagement and technology decision-making support.

Healthcare professionals expressed enhanced confidence in technology discussions, with one participant stating: *“This is a very educational course that will help me, and many others better understand the new technologies incorporating AI in the hospitals*.*”* Such feedback indicates successful preparation for organizational AI adoption and technology evaluation processes.

Faculty participants demonstrated curriculum integration intentions, with responses suggesting immediate application to educational practice. The high faculty satisfaction and commitment scores (Table 3) align with qualitative evidence of curriculum development and student preparation activities. Several faculty participants reported plans to incorporate course content into existing nursing informatics and technology courses, indicating cascading educational impact beyond direct program participation.

Organizational leaders reported improved staff preparedness for AI technology discussions and enhanced capacity for informed decision-making regarding technology adoption. Healthcare institutions noted increased staff engagement in AI-related policy and procedure development, suggesting successful workforce preparation for technology implementation processes.

Early evidence suggests successful bridging of academic learning with practical application, as participants translate course content to real-world healthcare challenges. The combination of high quantitative outcomes and positive qualitative feedback indicates effective achievement of program objectives for both individual professional development and organizational capacity building in healthcare AI adoption.

The program’s impact extends beyond individual learning to organizational readiness and professional networking. Participants reported forming professional connections focused on AI implementation, creating communities of practice that support ongoing learning and technology adoption. This network effect amplifies individual learning outcomes through peer support and knowledge sharing mechanisms.

## Discussion and Implications

### Innovation Contributions to Healthcare Education

This micro-learning AI education program demonstrates significant pedagogical advances addressing longstanding challenges in healthcare professional development. The measured knowledge gains represent meaningful learning achievement that exceeds typical effect sizes reported for traditional continuing education interventions in healthcare. With participants across two institutional partners achieving consistently high satisfaction scores (4.58-4.67), the program validates micro-learning as an effective approach for sophisticated technical education in healthcare settings.

The industry-academia partnership model proved essential for maintaining curriculum relevance while preserving educational rigor. Content relevance scores consistently ranked highest among all measures (4.61-4.71 across student and faculty groups), indicating successful integration of academic pedagogical standards with industry practical requirements. This achievement addresses a critical gap in healthcare AI education literature, where programs typically emphasize either theoretical knowledge or practical application but rarely achieve effective integration of both domains.

Participant testimonials demonstrate meaningful translation of learning to professional practice. One participant noted sharing information with their Chief for Virtual Health as they navigate AI within their facility, illustrating direct organizational impact through leadership engagement. Another participant emphasized how the course helps “many others better understand the new technologies incorporating AI in the hospitals,” indicating successful preparation for organizational AI adoption and technology evaluation processes.

### Addressing Critical Educational Gaps

The program addresses multiple critical gaps identified in healthcare AI education through targeted curriculum design and delivery innovation. As the first comprehensive, nursing-specific AI education program with demonstrated scale and effectiveness, it fills a significant void in professional development offerings. The strong participation by registered nurses (49% of completions) and measurable knowledge gains within this group (0.64 points) directly address workforce preparation needs highlighted in nursing literature.

The curriculum successfully bridges theoretical AI knowledge with practical healthcare implementation through progressive complexity design and authentic case studies. Participants’ high ratings for ability to integrate learning (4.53-4.57) and intention to apply skills (4.54) indicate effective translation of abstract concepts to actionable competencies. Faculty participants demonstrated exceptional outcomes across all dimensions, with 99% agreeing that course outcomes were met and 99% expressing commitment to applying what they learned, creating multiplier effects for ongoing workforce development.

The flexible delivery model accommodates diverse learner needs through self-paced, asynchronous access combined with interactive elements. The program’s success with over 3,000 participants across multiple institutions demonstrates financial sustainability while maintaining educational quality, addressing resource constraints that traditionally limit healthcare organizations’ investment in comprehensive technology training programs.

### Implications for Nursing and Healthcare Organizations

#### Workforce Development Strategy

The program provides evidence-based foundation for preparing nursing workforce for AI-enhanced healthcare delivery through systematic competency development. The significant knowledge gains (F(1, 1085) = 119.51, p < 0.001) and high commitment to application scores demonstrate effective workforce preparation extending beyond awareness to implementation readiness. Healthcare organizations can leverage this educational model to support strategic technology adoption while ensuring staff preparation and buy-in.

The scalable model offers organizational framework for technology adoption support and change management through structured educational intervention. Faculty satisfaction scores (4.67) and strong curriculum integration intentions can create multiplier effects, enabling ongoing workforce development capacity within organizations. This educational infrastructure may support sustained technology adoption rather than one-time training events.

Registered nurses demonstrated the largest knowledge gains among professional groups, positioning nursing professionals for expanded roles in organizational technology initiatives and policy development. The program’s emphasis on nursing-specific applications addresses the unique professional requirements and scope of practice considerations essential for nursing leadership in AI adoption.

#### Organizational Readiness and Quality Implications

Staff education serves as critical component of successful AI technology adoption through improved understanding of capabilities, limitations, and implementation requirements. The measured knowledge improvements and high content relevance scores (4.61-4.71) indicate effective preparation for informed technology evaluation and implementation decisions. Organizations can use educational outcomes to assess readiness for specific AI technology deployments.

Enhanced organizational capacity for technology evaluation and implementation decisions results from improved AI literacy across professional roles. The broad participation across healthcare disciplines creates interdisciplinary competency foundation essential for collaborative technology adoption. As one participant noted, the course provides “valuable evidence-based information that was easy to follow and understand,” supporting informed decision-making processes.

### Future Directions and Scalability

Expansion opportunities include application to other healthcare professions through adapted content while maintaining core pedagogical principles. The successful participation by diverse healthcare professionals demonstrates broader applicability beyond nursing. International expansion potential exists through cultural and regulatory adaptations while maintaining evidence-based instructional design principles.

The convergence of urgent AI education needs and proven microlearning effectiveness positions this model as essential for systematic workforce preparation as healthcare continues evolving. As one participant observed, “The world is evolving every day, the use of AI is also increasing.

Sharing knowledge about it is very important.” The program provides scalable framework for addressing ongoing technology education challenges while maintaining educational quality and professional standards.

## Conclusion

This micro-learning AI education program represents a shift in healthcare professional development, demonstrating that complex technical content can be effectively delivered through bite-sized, accessible modules while achieving meaningful learning outcomes. The partnership between Chamberlain University, Walden University and Hippocratic AI successfully bridges academic expertise and industry relevance, creating a scalable model for addressing rapid technological advancement in healthcare education.

The program’s success with over 3,000 participants validates micro-learning as a viable solution to the urgent need for AI literacy in nursing and healthcare. The significant knowledge gains and consistently high satisfaction scores (4.58-4.67) demonstrate that working professionals can effectively acquire sophisticated AI competencies through flexible, self-paced learning. The particularly strong outcomes among registered nurses, who showed the largest knowledge gains (0.64 points), position the nursing profession to lead healthcare technology transformation. The emphasis on nursing-specific applications through dedicated AINP courses addresses a critical gap in AI education, providing content that directly relates to nursing scope of practice and professional leadership opportunities. Faculty development outcomes demonstrate dual impact on immediate professional preparation and long-term educational capacity building, with 99% of faculty participants expressing commitment to applying their learning, creating cascading effects that extend beyond direct enrollment.

This program provides a replicable framework for addressing emerging technology education needs across healthcare disciplines. The template approach and shared resource model enable rapid scaling while maintaining content quality, creating opportunities for broader workforce development initiatives. As AI technologies continue evolving throughout healthcare delivery, the proven effectiveness of this micro-learning approach positions it as an essential component of strategic workforce preparation.

The program’s success challenges traditional assumptions about continuing education delivery and demonstrates that innovative partnerships between academia and industry can produce educational solutions meeting the complex demands of modern healthcare practice. By combining pedagogical expertise with technological insights, this model offers a blueprint for preparing healthcare professionals to lead AI implementation and ensure ethical, safe, and effective integration into patient care.

## Data Availability

Data supporting the results can be accessed by contacting the corresponding author.

